# Novel Insights for Radiation Risk Assessment Unveiled by Deep Learning

**DOI:** 10.1101/2024.04.27.24306487

**Authors:** Zhenqiu Liu, Igor Shuryak, David J Brenner, Robert L Ullrich

## Abstract

Contemporary radiation risk assessment predominantly depends on nonlinear parametric models, which typically include a baseline term, a dose-response term, and an effect modifier term. Despite their widespread application in estimating tumor risks, parametric models face a notable drawback: their rigid model structure can be overly restrictive, potentially introducing bias and inaccuracies into risk estimations.

In this study, we analyze data on solid tumors and leukemia from the Life Span Study (LSS) to compare the performance of deep neural network (DNN) and nonlinear parametric (NLP) models in assessing ERRs. DNN presents novel perspectives for radiation risk assessment. Our findings indicate that DNN can perform better than the traditional parametric models. Even if DNN and NLP models exhibit similar performance in predicting tumor incidence, they diverge significantly in their estimated ERRs. Standard NLP models tend to underestimate ERRs directly linked to radiation dose, overestimate ERRs for individuals at younger attained ages and ages at exposure, and underestimate ERRs for those at older attained ages. Furthermore, DNN consistently identifies radiation dose as the primary and predominant risk factor for ERRs in leukemia and solid tumors, underscoring the critical role of radiation dose in risk assessment. The insights from DNN could enhance low-dose radiation risk assessment and improve parametric model development.

## Introduction

Accurately estimating radiation risk is paramount for safeguarding public health, establishing robust safety standards, and fostering advancements in radiation safety research. In this pursuit, Life Span Studies (LSS) involving A-bomb survivors from Hiroshima and Nagasaki have served as a crucial foundation for radiation risk assessment. The LSS cohort comprises more than 86,000 survivors located within 10 km of the bomb epicenters, offering a substantial and diverse population encompassing all age groups and both genders, with an extensive follow-up time. This expansive and thorough cohort stands as the gold standard for radiation risk assessment [1-5]. We will utilize the publicly available summary tables derived from this cohort for our study [6-7], which include the solid tumor incidences from 1958-1998 and leukemia incidences from 1950-2001.

To date, radiation risk assessments have typically relied on a parametric model in the following functional structure:

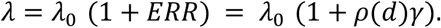

Where *λ* represents the expected tumor incidence rate, *λ*_0_ is the baseline incidence rate with zero dose, but may depend on attained age, age at exposure, time since exposure, sex, city, and other factors; *ρ*(*d*) describes the shape of dose-response including linear dose (*ρ*(*d*) = *β*_1_*d*) and linear-quadratic (*ρ*(*d*) = *β*_1_*d* + *β*_2_*d*^2^); and *γ* represents the effect modifier associated with the effect of dose. It describes how the level of radiation-related ERR varies with city, gender, age at exposure, attained age, and time since exposure. While the model incorporates slightly varying risk factors, it employs a standardized parametric structure to evaluate radiation risks across diverse tumors. Although this model is straightforward and facilitates interpretation, its inherent rigidity may limit its ability to encompass the intricate dose-response relationships present in the data fully. The specifics of its formulation significantly influence the effectiveness of a parametric model. Model misspecification has the potential to introduce bias and inaccuracies in risk estimations.

Deep learning, or deep neural networks (DNN), stands at the forefront of modern AI, offering unparalleled flexibility and data-driven capabilities. It has demonstrated state-of-the-art performance across various domains, breaking through limitations imposed by traditional function forms [8-12]. However, despite these advancements, the full scope of their potential, implications, and limitations in radiation risk assessment remains unexplored.

In this paper, we propose the application of the Poisson DNN model to publicly available solid tumor and leukemia data. Our objective is to assess the excess relative risk (ERR) associated with solid tumors and leukemia, pinpoint the risk factors associated with each cancer, and investigate the potential of DNN in radiation risk estimation. Through this exploration, we aim to contribute valuable insights to the understanding of deep learning’s role in enhancing radiation risk assessment methodologies.

## Methods

### Datasets

#### Solid Tumor Incidence Data

The person-year summary data (LSSinc07) [6] were obtained from the Radiation Effects Research Foundation (RERF) website (https://www.rerf.or.jp/en/). After excluding records with missing doses, we identified 25,570 cells containing summarized data from 17,448 solid tumors among 105,427 survivors, with a total of 2,764,735 person-year follow-ups. The dataset comprises 59.3% women and 40.7% men, with 69.7% residing in Hiroshima, 30.3% in Nagasaki, and 23.9% classified as NIC.

#### Leukemia Incidence Data

The data for this study was retrieved from the RERF website [7]. The person-year summary table contains 36,841 cells after excluding entries with missing doses. The dataset encompasses 113,011 subjects, among whom there were 371 incidences of leukemia. The distribution of gender in the dataset is 58.6% women and 41.4% men with 69.5% from Hiroshima, 30.5% from Nagasaki, and 23.45% classified as NIC.

### Deep Learning Models

We employed a deep neural network with identical architecture for predicting radiation risk in both solid tumor and leukemia incidence data, as illustrated in Figure 1.

**Figure 1:**
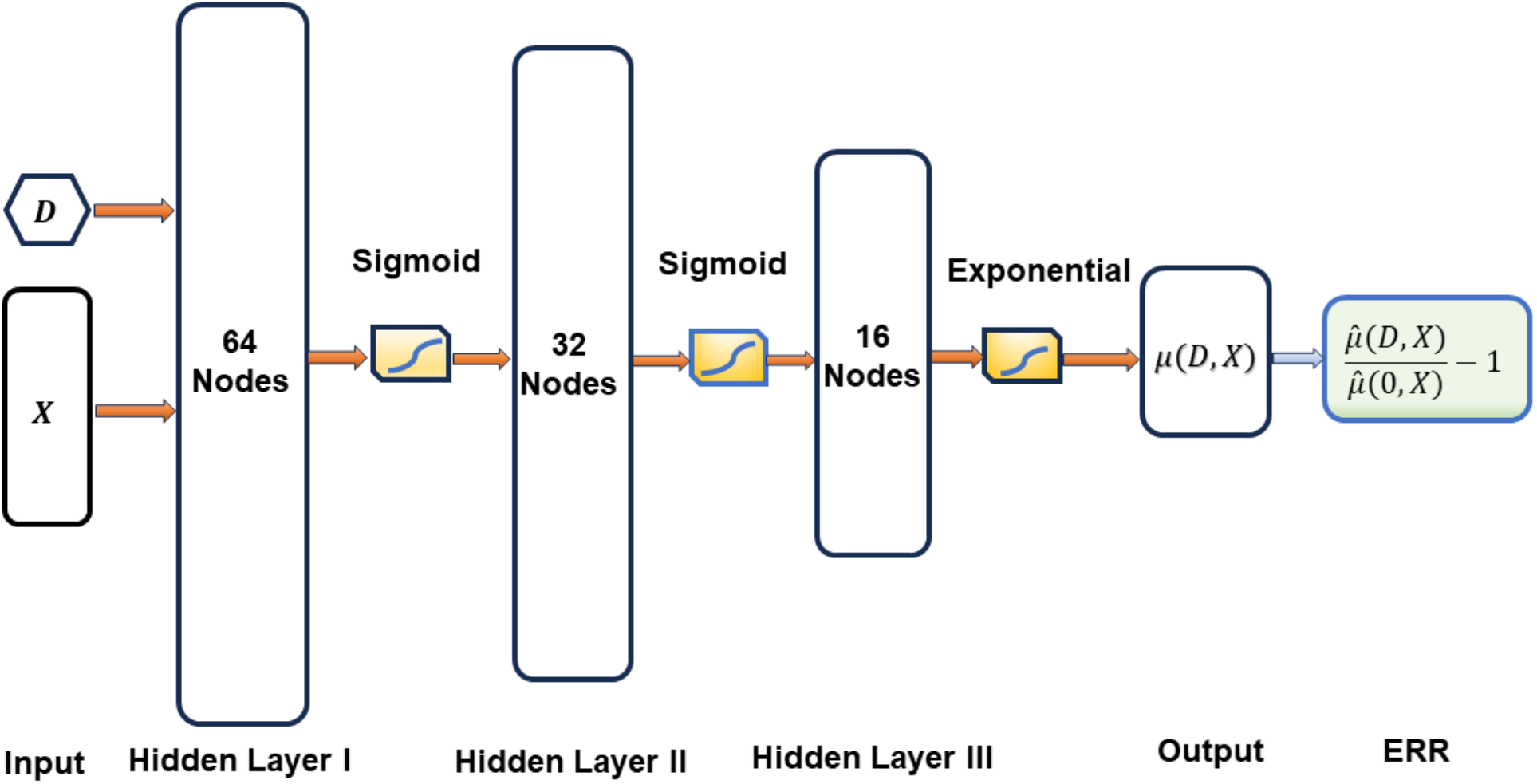
Neural network architecture for radiation risk prediction, with D representing radiation dose and X representing confounding factors. ERR denotes excess relative risk. The network comprises three hidden layers with 64, 32, and 16 nodes, respectively. Both sigmoid and exponential activation functions were employed. The rightmost shaded box represents the estimated ERR after training the model.

Given radiation dose (D), other associated factors (X), cancer incidence (Y), and person-year (Py), we establish a neural network with three hidden layers, each consisting of 64, 32, and 16 nodes, respectively. With the assumption of Poisson distribution, for each cell i,

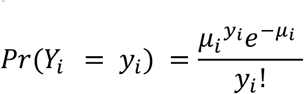

and

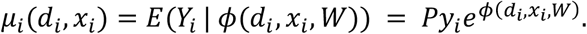

The loss function is the negative log-likelihood (NLL) of a Poisson distribution as

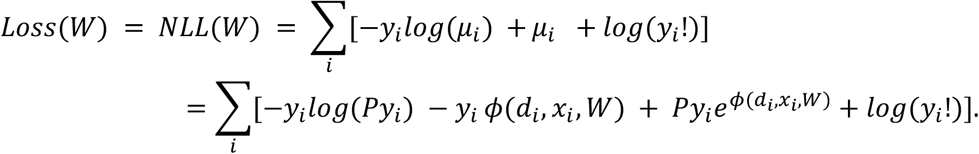

The software is implemented in Python and TensorFlow. After training the model, the excess relative risk (ERR) is estimated as

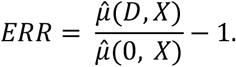

#### Hyperparameter Determination and Model Comparison

For the sake of comparison, we opted for identical numbers of hidden layers and nodes in both solid tumor and leukemia analyses. The determination of the number of hidden layers and nodes is guided by minimizing the negative log likelihood of the test data through cross-validation. A learning rate of 0.0001 is set, and the widely used Adam optimization algorithm is employed for parameter estimation. The Adam algorithm harnesses adaptive learning rates, determining individual rates for each parameter. The actual learning rate of Adam in each iteration is confined by the step size hyperparameter (0.0001).

The comparison of various models is conducted using Shapley Additive Explanations (SHAP) values [13-15]. SHAP is a method that elucidates individual predictions based on the Shapley values derived from game theory. Shapley values conceptualize a prediction as a game played by the feature values, comparing the prediction of each feature with its average prediction. These values guide us on how to equitably distribute the “payout” (i.e., the prediction) among the features. In this analogy, a player represents an individual feature value, and the prediction stands as the payout. Shapley values break down a model’s predictions into contributions that can be additively attributed to different explanatory variables (features). This method provides a systematic approach for determining the contributions of features to individual predictions in any machine learning model.

## Results

### DNN achieves comparable or marginally better performance in predicting tumor incidence

We employed both a DNN and a nonlinear parametric (NLP) model for analyzing leukemia and solid tumor data. The DNN utilizes a standardized network architecture for both leukemia and solid tumor prediction, consisting of 3 hidden layers with 64, 32, and 16 nodes respectively. Nonlinear parametric models, estimated by GAM R package, represent some of the most effective models for radiation risk assessment in the literature [6-7]. The specific nonlinear parametric model for leukemia is estimated as follows:

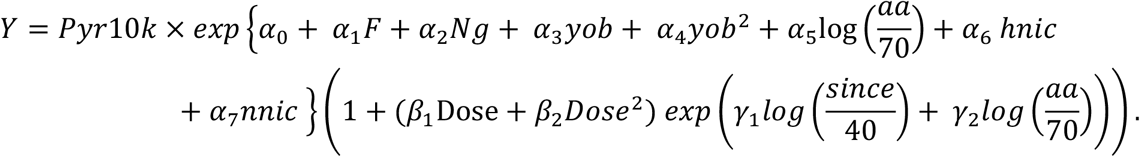

Where Pyr10k: *Pyr*/10000; F: female; Ng: Nagasaki; yob: year of birth; hnic: *Hiroshima* × *nic;* nnic: *Nagasaki* × *nic;* nic: not in the city; aa: attained age; and since: time since exposure. Similarly, the nonlinear parametric model for solid tumor is defined as follows:

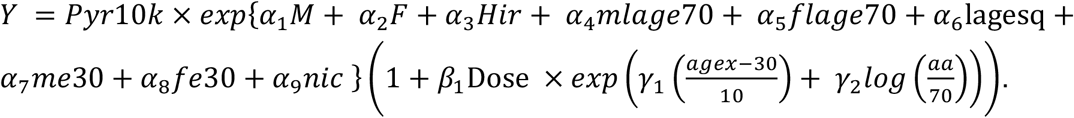

Where M: male; F: female; Hir: Hiroshima; aa: attained age; agex: age at exposure; mlage70: 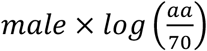; flage70: 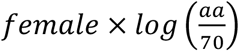; lage70sq: 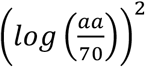; me30: 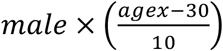; fe30: 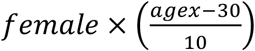; and nic: not in the city.

We employed a 5-fold cross-validation to evaluate the model performance for both leukemia and solid tumors. We divided the datasets into 5 equal-sized subsets, trained the model with four subsets, and assessed its performance with the remaining subset. This process was repeated 5 times, and the final performance metric is the average. For the DNN model, an additional 10% of the training data is used as a validation set to control the model complexity. The results are reported in Table 1:

**Table 1:** Performance comparison of the DNN and nonlinear parametric (NLP) model with solid leukemia and solid tumor data, where MNLL: mean negative log-likelihood; MPDL: mean Poisson deviance loss; MPR: mean Pearson residuals; MAE: mean absolute error; and RMSE: root mean square error. A bolded smaller number denotes better performance.

| Tumors | Models | MNLL | MPDL | MPR | MAE | RMSE |
| --- | --- | --- | --- | --- | --- | --- |
| Leukemia | NLP | 0.044 | 0.070 | 0.152 | 0.019 | 0.095 |
|  | DNN | 0.044 | 0.070 | **0.151** | **0.018** | 0.095 |
| Solid Tumors | NLP | 0.619 | 0.587 | 0.567 | 0.413 | 0.849 |
|  | DNN | **0.618** | **0.585** | **0.566** | **0.409** | **0.839** |

As depicted in Table 1, the deep neural network (DNN) with 64, 32, and 16 nodes for 3 hidden layers achieves comparable performance in predicting leukemia incidences across various metrics. Specifically, the DNN exhibits test Mean Negative Log-Likelihood (MNLL), Mean Poisson deviance Loss (MPDL), Mean Pearson Residuals (MPR), Mean Absolute Error (MAE), and Root Mean Squared Error (RMSE) of 0.044, 0.070, 0.151, 0.018, and 0.095, respectively, which closely aligns with the NLP model’s performance across different metrics. Moreover, the DNN demonstrates marginally better performance in predicting solid tumor incidence. It registers test MNLL, MPDL, MPR, MAE, and RMSE values of 0.618, 0.585, 0.566, 0.409, and 0.839, respectively, which outperforms the NLP model with corresponding values of 0.619, 0.587, 0.567, 0.413, and 0.849, respectively.

However, it’s important to note that there may exist superior nonlinear parametric and DNN models for predicting tumor incidence. Our deliberate choice of a model with similar performance aims to demonstrate that even with comparable predictive capabilities for tumor incidence, the excess relative risk (ERR) can exhibit significant variation between DNN and NLP models.

### The SHAP values for tumor incidences with both DNN and NLP highlight the distinct risk factors associated with solid tumors and leukemia

We assess the significance of each feature in predicting tumor incidence using SHAP values for both leukemia and solid tumors. Computational findings are presented in Figure 2.

**Figure 2:**
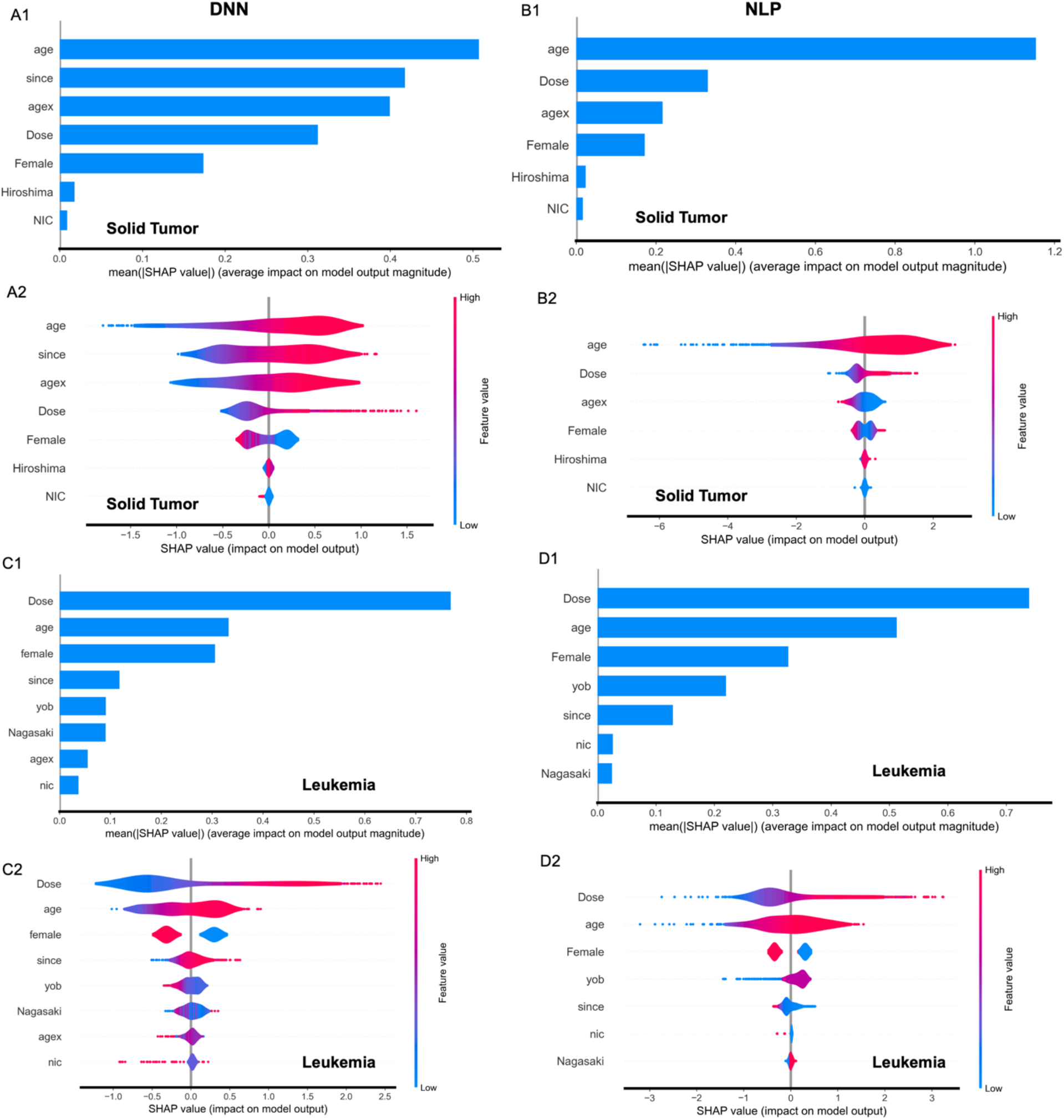
SHAP values for predicting cancer incidence of solid tumors and leukemia. The top four panels, A1-A2 and B1-B2, illustrate the results for solid tumors from DNN and NLP, respectively. The bottom four panels, C1-C2 and D1-D2, depict the results for leukemia from DNN and NLP, respectively.

As illustrated in the top four panels of Figure 2, the top five risk factors identified by DNN (Figure 2A1-2A2) for predicting solid tumor incidence are attained age (age), time since exposure (since), age at exposure (agex), radiation dose, and sex. Remarkably, these features closely resemble the top four risk factors revealed by NLP (Figure 2B1-2B2) for solid tumor incidence prediction, which consist of attained age, radiation dose, age at exposure, and sex. It’s worth noting that the features attained age, age at exposure, and time since exposure are entirely collinear. While DNN permits the inclusion of all three features in the model, NLP does not. However, incorporating these collinear features doesn’t enhance prediction accuracy. Nevertheless, it enables us to dissect the individual contributions of each feature—an advantage unique to DNN.

Comparable findings for leukemia are depicted in the lower four panels of Figure 2. The top five risk factors identified by DNN (Figure 2C1-2C2) for forecasting leukemia incidence are radiation dose, attained age, sex, time since exposure, and year of birth (yob). Remarkably, these align precisely with the top five risk factors identified by NLP (Figure 2D1-2D2), albeit with slight variations in rank order.

Take a closer look at the violin plots in Figure 4 (A2, B2); there are notable distinctions between DNN and NLP. DNN revealed a positive correlation between age at exposure and the SHAP value of solid tumor incidence, whereas NLP suggested a negative association between age at exposure and the SHAP values of solid tumor incidence. However, both DNN and NLP identified attained age as a primary risk factor for solid tumors and radiation dose as the primary risk factor for leukemia. This underscores the biological and clinical disparities between solid tumors and leukemia.

**Figure 3:**
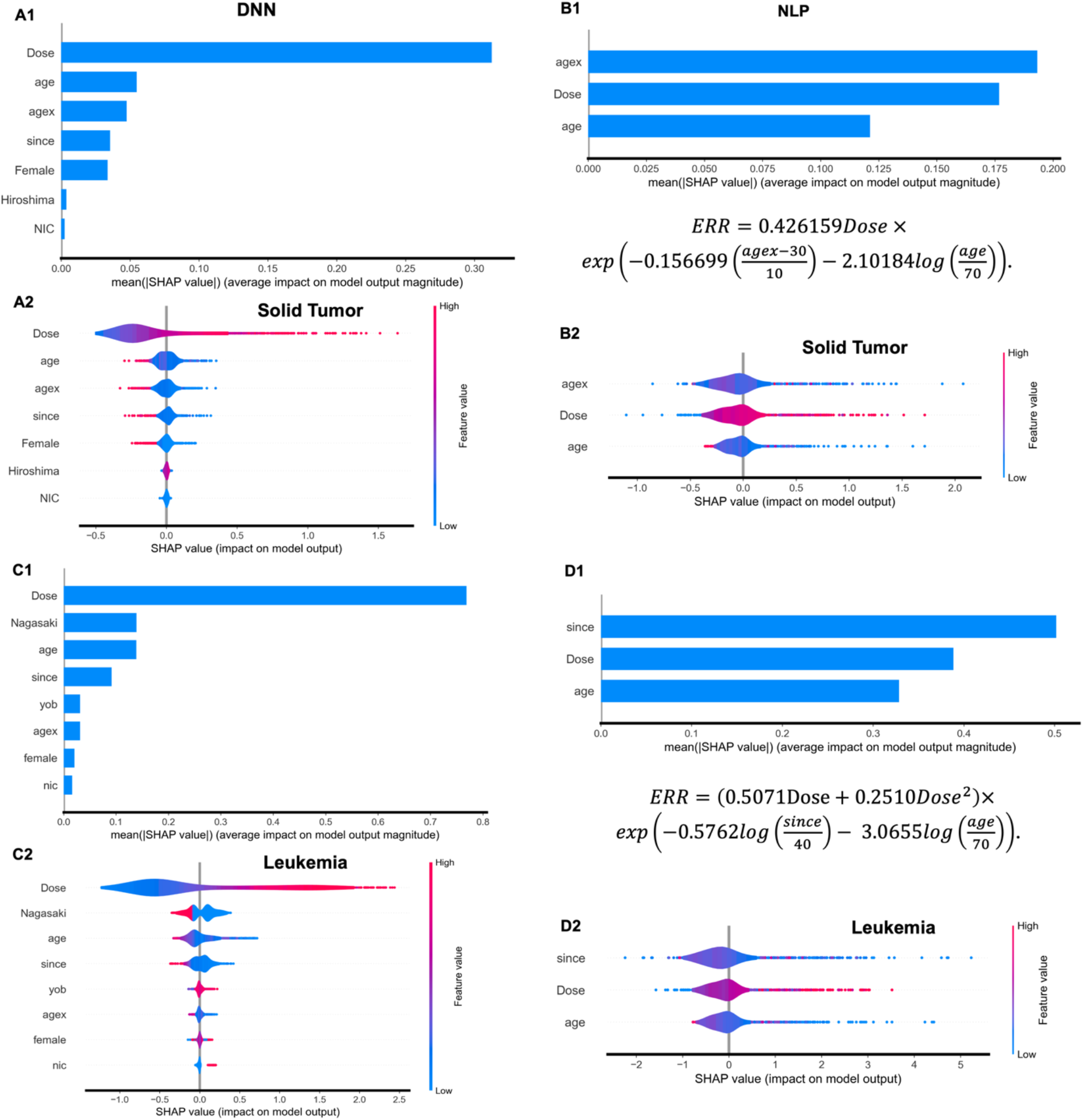
SHAP values for predicting ERRs of solid tumors and leukemia. The top four panels, A1-A2 and B1-B2, illustrate the results for solid tumors from DNN and NLP, respectively. The bottom four panels, C1-C2 and D1-D2, depict the results for leukemia from DNN and NLP, respectively.

**Figure 4.**
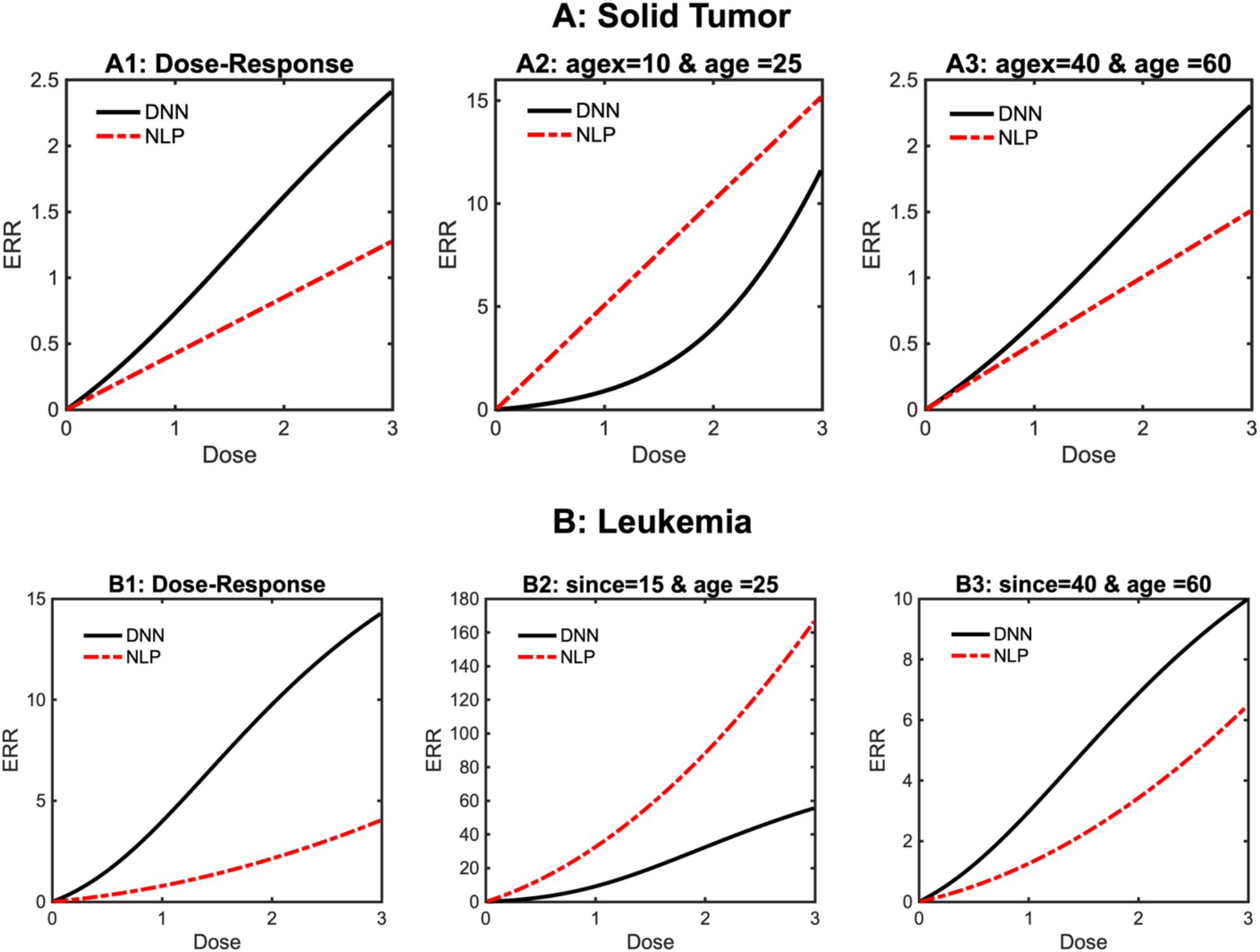
depicts the dose-response relationships of DNN and NLP for solid tumors (top panels: A1-A3) and leukemia (bottom panels: B1-B3). Panels A1 and B1 illustrate the dose-response relationships after excluding the effects of other features, while panels A2-A3 and B2-B3 showcase the dose-response curves across different age groups.

### The SHAP values for ERRs of solid tumor and leukemia highlight the difference between DNN and NLP

We delve deeper into the risk factors linked with excess relative risks (ERRs) of solid tumors and leukemia using SHAP values for ERR. The computational findings are detailed in Figure 3.

Comparing the SHAP values of ERRs for solid tumors between DNN (A1-A2) and NLP (B1-B2), DNN identified radiation dose as the top and most significant risk factor associated with ERRs of solid tumors, with a mean absolute SHAP value of 0.34. In contrast, radiation dose ranked second among the risk factors associated with ERRs of solid tumors in the NLP model. The mean absolute SHAP values of ERRs for radiation dose by the NLP model are around 0.18, significantly lower than those from DNN (0.34). This suggests that NLP might underestimate the ERRs directly determined by radiation dose. Conversely, NLP (B1-B2) identified age at exposure and attained age as the top and third risk factors for ERRs of solid tumors, with mean absolute SHAP values of 0.20 and 0.12, respectively. These values are notably higher than the mean absolute SHAP values of age at exposure and attained age (0.04 and 0.05) determined by DNN (A1-A2). This suggests that NLP may overestimate ERRs attributed to age-related factors. It’s noteworthy that while age at exposure ranks third in predicting tumor incidence (Figure 2B1-2B2) within NLP, it emerges as the top risk factor for ERRs, which appears counterintuitive.

Comparable findings for leukemia are illustrated in the lower four panels of Figure 3. The DNN (C1-C2) identified radiation dose as the primary and most significant risk factor associated with the ERRs of leukemia, with a mean absolute SHAP value of approximately 0.8. This value is notably higher than the mean absolute SHAP value of ERR (0.4) determined by NLP (D1-D2), where radiation dose ranks as the second significant risk factor associated with ERRs within the NLP model. This implies that NLP may potentially underestimate the ERRs directly attributed to radiation dose. Conversely, NLP (D1-D2) identified time since exposure as the primary and attained age as the third most significant risk factors for excess relative risks (ERRs) of leukemia, with mean absolute SHAP values of 0.50 and 0.33, respectively. These values are significantly higher than the mean absolute SHAP values of time since exposure and attained age (0.08 and 0.12) determined by DNN (D1-D2). This implies that NLP might overestimate ERRs attributed to age-related factors. There are also inconsistencies between the identified risk factors for tumor incidence and excess relative risks (ERRs) in NLP. While radiation dose emerges as the primary risk factor for leukemia incidence (Figure 2D1-2D2), it ranks as the second significant risk factor for ERRs. Additionally, the time since exposure, which is only the fifth significant risk factor for tumor incidence, becomes the primary risk factor for ERRs. This reversal is counterintuitive.

### The dose-response curves generated by DNN and NLP underscore certain limitations of NLP models

We further explore the dose-response relationships across different age groups for both solid tumor and leukemia, and the results for DNN and NLP are reported in Figure 4 and Supplementary Table 1.

As illustrated in Figure 4, panels 4A1 and 4B1 display the dose-response relationships after mitigating the effects of other features. In the DNN model, we achieve this by setting other features (covariates) to their mean values, as we conducted z-score normalization for all variables using a normalization layer in the DNN architecture. For the NLP model, we neutralize the influence of age modifiers by standardizing the age at exposure to 30 and the attained age to 70 for solid tumors, and setting the time since exposure to 40 with an attained age of 70 for leukemia. Figure 4A1 and 4B1 distinctly show that NLP underestimated the excess relative risks (ERRs) of radiation dose compared to DNN for both solid tumors and leukemia. This observation aligns with the patterns observed when comparing Figures 2B1-2B2 and 2D1-2D2 to Figures 3B1-3B2 and 3D1-3D2 in earlier sections. Specifically, the SHAP values corresponding to radiation doses for ERRs were approximately halved compared to those for tumor incidences.

Both Figures 4A2-4A3 and 4B2-4B3 and Supplementary Figure 1 and Table 1 reveal that, in comparison to DNN, NLP overestimated the excess relative risks (ERRs) for subjects with younger attained ages, while underestimating the ERRs for subjects with older attained ages for both leukemia and solid tumors. The model structure of ERR in NLP may contribute partly to these discrepancies. Specifically, ERR is defined as ρ(d)γ, where for solid tumors:

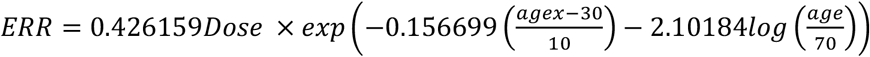

with a linear dose-response ρ(*d*) = 0.426159*Dose* and an age modifier of 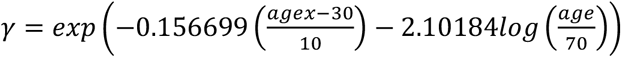.

For leukemia:

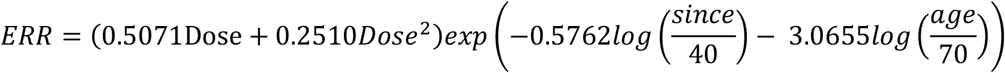

with a linear-quadratic dose respose ρ(*d*) = (0.5071*Dose* + 0.2510*Dose*^2^) and an age modifier of 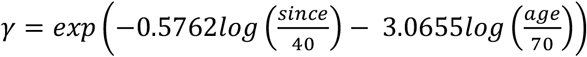.

This model specification results in ERRs being heavily influenced by the age modifiers, which may introduce bias and inaccuracies in risk assessments.

## Discussion and Conclusions

The popular approach for radiation risk assessment involves a nonlinear parametric (NLP) Poisson model, expressed as λ_0_ (1 + ρ(*d*)γ). The parameters are typically estimated using maximum log-likelihood of Poisson regression, with the ERR given by *ERR* = ρ(*d*)γ being estimated indirectly during this process. This framework is straightforward and is widely utilized by both academics and governmental agencies. However, various parametric models can achieve comparable performance in predicting tumor incidences, but they may diverge significantly in ERR prediction. Selecting an appropriate model structure for ERRs is crucial yet challenging for accurately estimating radiation risk within a parametric model.

In this paper, we compare radiation risk assessments using both a DNN and an NLP model with solid tumor and leukemia data. The data-driven DNN model is highly flexible and capable of approximating underlying functions effectively with a sufficient number of layers and nodes. Unlike parametric models, it mitigates the inherent risk of model misspecification. We discover that despite achieving similar performance in tumor incidence prediction between DNN and NLP, the NLP model, with its commonly used structure, tends to underestimate the direct impact of radiation dose on ERR. Furthermore, it overestimates the ERRs for subjects with younger attained ages and ages at exposures, while underestimating the ERRs for subjects with older attained ages. These findings underscore certain potential limitations of widely adopted parametric models.

Despite its computational complexity and black-box nature, DNN offers unique perspectives for radiation risk assessment. Specifically, DNN yields higher estimates of ERRs compared to those obtained from NLP models. Additionally, age factors exert less influence on ERRs in DNN. Furthermore, DNN consistently identifies radiation dose as the primary and predominant risk factor for ERRs of leukemia and solid tumors, highlighting the crucial significance of radiation dose in risk assessment. These novel insights from DNN can be used for low-dose radiation risk assessment and better parametric model construction.

## Data Availability

All data produced in the present study are available upon reasonable request to the authors

## ACKNOWLEDGMENTS

The Radiation Effects Research Foundation (RERF), Hiroshima and Nagasaki, Japan is a private, nonprofit foundation funded by the Japanese Ministry of Health, Labour and Welfare (MHLW) and the U.S. Department of Energy (DOE). This research was also funded in part through the DOE award DE-HS0000031 to the National Academy of Sciences (ZL). The views of the authors do not necessarily reflect those of the two governments.

---

**Supplementary Table 1:** Comparison of ERR Estimates for DNN and NLP Models Across Different Ages at Exposure (Agx) and Attained Ages (Age) for Solid Tumors and Leukemia. The values represent the mean ERRs within the specified age at exposure and attained age group, alongside their corresponding dose ranges. Standard errors (SE) are provided in parentheses.

| ERR | Dose (Gy) | Agx 0-10 & Age: 20-40 |  | Agx: 35-45 & Age:60-75 |  |
| --- | --- | --- | --- | --- | --- |
|  |  | DNN | NLP | DNN | NLP |
| Solid Tumor | 0-0.25 | 0.0553<br>(±0.0015) | **0.3743**<br>**(±0.0113)** | **0.0401**<br>**(±0.0013)** | 0.0341<br>(±0.0011) |
|  | 0.25-0.5 | 0.2307<br>(±0.0054) | **1.4274**<br>**(±0.0496)** | **0.1651**<br>**(±0.0045)** | 0.1364<br>(±0.0027) |
|  | 0.5-1.0 | 0.645<br>(±0.0157) | **3.1481**<br>**(±0.1105)** | **0.3635**<br>**(±0.0093)** | 0.2889<br>(±0.0057) |
|  | 1.0-2.0 | 1.9941<br>(±0.0513) | **6.285**<br>**(±0.1745)** | **0.7675**<br>**(±0.0174)** | 0.5871<br>(±0.0096) |
|  | 2.0-3.0 | 6.3879<br>(±0.1986) | **10.9444**<br>**(±0.3398)** | **1.2741**<br>**(±0.0313)** | 0.9764<br>(±0.0143) |
|  | >3.0 | 10.6032<br>(±0.6349) | **13.6183**<br>**(±0.7841)** | NA | NA |
| Leukemia | 0-0.25 | 0.2704<br>(±0.0072) | **1.4717**<br>**(±0.0604)** | **0.1765**<br>**(±0.0057)** | 0.0709<br>(±0.0025) |
|  | 0.25-0.5 | 1.2865<br>(±0.0236) | **6.2276**<br>**(±0.3947)** | **0.8071**<br>**(±0.0158)** | 0.3211<br>(±0.0107) |
|  | 0.5-1.0 | 4.2327<br>(±0.0998) | **16.2597**<br>**(±1.0169)** | **2.2073**<br>**(±0.0560)** | 0.8369<br>(±0.0289) |
|  | 1.0-2.0 | 13.4548<br>(±0.3082) | **39.7454**<br>**(±1.9251)** | **5.2906**<br>**(±0.1139)** | 2.0595<br>(±0.0569) |
|  | 2.0-3.0 | 29.9668<br>(±0.9052) | **88.8133**<br>**(±6.2834)** | **9.7351**<br>**(±0.2340)** | 4.9597<br>(±0.1592) |
|  | >3.0 | 39.0851<br>(±1.1946) | **139.2663**<br>**(±9.5777)** | **11.3917**<br>**(±0.4486)** | 6.4541<br>(±0.3144) |

**Supplementary Figure 1:**
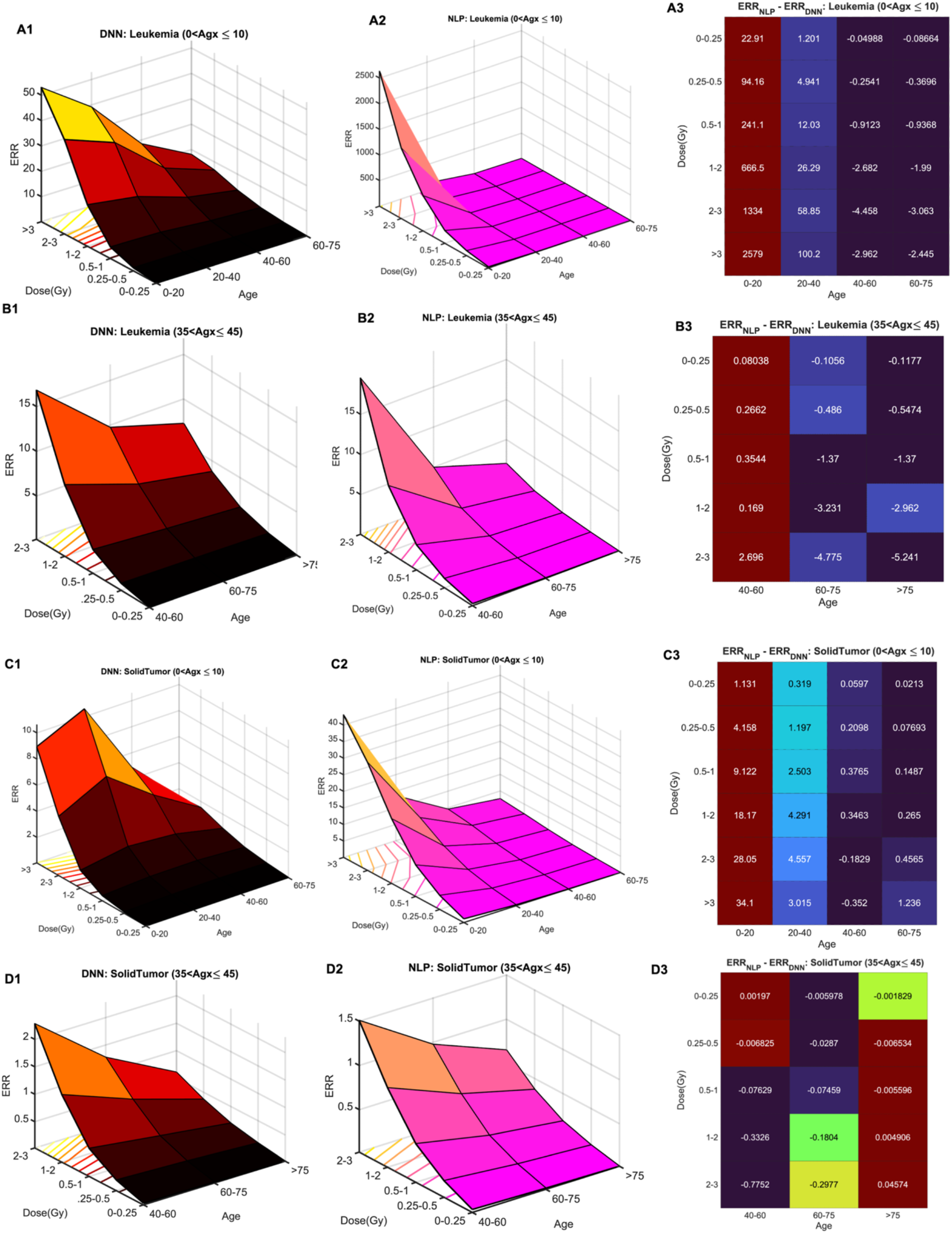
ERRs estimated at different age at exposure (Agx), attained age (Age), and radiation dose groups. A1-3-B1-3: Leukemia; C1-3-D1-3: Solid Tumor. NLP: Nonlinear Parametric Model; DNN: Deep neural network.

